# Factors Associated with All-cause Mortality and Morbidity of Motorcycle Crash-Related Neurological and Musculoskeletal Injuries: The MOTOR Trial Ancillary Study

**DOI:** 10.1101/2025.02.21.25322634

**Authors:** Herman Lule, Micheal Mugerwa, Anne Abio, Benson Oguttu, Andrew Kakeeto, Daniel Asiimwe, Harvé Monka Lekuya, Robinson Ssebuufu, Patrick Kyamanywa, Till Bärnighausen, Michael Lowery Wilson, Jussi P. Posti

## Abstract

**Introduction:** The objective of this study was to examine the factors linked to all-cause mortality and morbidity from neurological and musculoskeletal injuries in motorcycle accidents.

**Methods:** The study was part of a two-armed, parallel, multi-period, cluster-randomized controlled trial of 1003 motorcycle crash victims. Morbidity was assessed using various scoring systems, and mixed effects regression models were employed for analysis.

**Results:** 90-day all-cause mortality was 9.2% (82/887). Factors associated with mortality included referral-to-discharge >1 hour [OR 4.215 (1.802-9.858), *p*=.001), Kampala Trauma Score (KTS) ≤6 [OR 7.696 (1.932-30.653), *p*=.004], GCS 9-12 [OR 3.432 (1.194-9.870), *p*=.022], GCS ≤8 [OR 6.919 (2.212-21.645), *p*=.001], intra-axial lesions [OR 78.647 (9.871-626.587), *p*<.001], extra-axial lesions [OR 11.933 (1.386-102.750), *p*=.024], skull fracture [OR 11.366, (1.197-107.977), *p*=.034], and decompressive craniotomy (DC) [OR 0.260 (.095-.706), *p*=.008).

A proportion of 14.5% had unfavourable Glasgow Outcome Scale (1-3); associated factors included increasing age [OR 1.02, (1.013-1.045, *p*<.001], multiple injuries [OR 4.559 (1.185-17.531), p=.027], KTS 7-8 [OR 2.755 (1.285-5.906), p=.009], KTS ≤6 [OR 7.551 (2.815-20.255), p=.001], GCS 9-12 [OR 4.07 (1.901-8.719), p=.001], GCS ≤8 [OR 13.779 (5.643-33.645), *p*<.001], and DC [OR 0.149 (.075-.295), *p*<.001].

Factors associated with unfavourable patient-reported musculoskeletal outcomes included being married [OR 1.984 (1.322–2.976), *p*=.001], multiple injuries [OR 1.762, (1.001–3.100), *p*=.049], and enrolment after the onset of the COVID-19 pandemic [OR 2.095 (1.199–3.659), p=.009].

**Conclusions:** The key determinants of mortality and adverse neurological and musculoskeletal injury outcomes observed in this study are essential for establishing core outcome sets in future research and predictive models.

**Summary of the Key Messages:**

- **Existing Knowledge:** Motorcycle crashes in low- and middle-income countries represent significant public health issues, exacerbated by low helmet usage and insufficient trauma care resources.
- **Study Contributions:** This MOTOR trial ancillary study investigated factors associated with all-cause mortality and morbidity from motorcycle crashes, emphasizing neurological and musculoskeletal injuries as critical targets for enhancing rural trauma care systems.
- **Implications for Research, Practice, and Policy:** Our findings indicate that the anatomical nature, multiplicity and severity of injuries, referral-dispatch intervals, operative interventions, and COVID-19-related social conditions significantly influence injury outcomes. These factors may serve as a foundational outcome set for benchmarking future clinical trials and trauma registries.

## INTRODUCTION

Motorcycle crashes represent a major global public health challenge, with associated injuries affecting both high-income and low-income countries (LICs) [1]. These accidents account for nearly half of emergency department admissions following road traffic incidents [2]. The resulting injuries often involve neurological damage or musculoskeletal trauma, with vulnerable road users, such as riders, passengers, and pedestrians, disproportionately impacted [3]. Traumatic brain injuries (TBIs) from motorcycle crashes lead to mortality rate of 3-6%, with higher rates among non-helmet users [4]. Survivors may experience substantial disability and financial burden [5].

In low-and-middle-income countries (LMICs), especially Africa, motorcycle crash-related injuries result in higher mortality and morbidity compared to high-income countries (HICs) [2], [3]. Factors include lower helmet usage (30.6% in Africa compared to a global average of 44.8%) [1], [2] and inadequate road safety legislation [6]. The continent’s high commercial motorcycle usage, driven by a favourable climate, exacerbates the problem. Additionally, limited access to post-crash care, due to a shortage of skilled healthcare workers and a weak prehospital system, worsens injury outcomes [7]. Africa has only 0.15 neurosurgeons per 100,000 people, far below the WHO recommendation [8].

Uganda, a low-income country, has a low helmet wearing rate of 19% [1] and faces a high burden of fatal motorcycle crashes, especially on rural low-capacity roads [9]. The inefficiency of the rural trauma care system, long transport distances, and ineffective referral protocols worsen injury outcomes [10]. Recognizing local factors affecting injury outcomes and barriers to post-crash care is essential for reducing negative impacts. Prior research has often combined all road traffic crash outcomes, neglecting the specificities of motorcycle incidents [11]. This study examined factors related to 90-day mortality and morbidity of neurological and musculoskeletal injuries in motorcycle accidents, which is crucial for improving rural trauma care in Uganda [12].

## METHODS

This study was nested within a larger cluster randomized trial which assessed the impact of rural trauma team development on outcomes of motorcycle-crash related injuries (Pan African Clinical Trial Registry: PACTR202308851460352). The parent trial was a two-armed, parallel, multi-period, cluster-randomized controlled trial of 1003 motorcycle crash victims (501 intervention, 502 control) enrolled at 3 intervention and 3 control trauma centres in Uganda [9]. Frontline care providers in the intervention group participated in the Rural Trauma Team Development Course of the American College of Surgeons, in conjunction with the establishment of rural trauma networks, in contrast to the control group [9]. Clusters were randomly assigned 1:1 using permuted block codes. Participants and outcome assessors were blinded to the allocation. The present ancillary study utilized patient-participant data on covariates collected from the motorcycle trauma outcome registry (MOTOR) project, which was executed in parallel to the trial [9]. The data were collected during August 2019 to August 2023, and the analyses were completed in Spring 2024.

### Study settings

The study was conducted in six level III trauma centres including Jinja, Hoima, Fort portal, Mubende, Kiryandongo and Kampala International University Teaching Hospitals. All referrals were handled at Mulago Hospital in Kampala, Uganda, which is the country’s comprehensive level I trauma centre. The level of specialized care and detailed patient work flow processes employed at these facilities have been comprehensively described in prior literature [9], [10].

### Eligibility criteria

The study population comprised patients aged 2 to 80 years who presented to six sites within 24 hours of a motorcycle crash-related injury. Pregnant women, mentally incapacitated individuals unable to consent, and stroke patients were excluded to limit reported morbidity to trauma.

### Sample size determination

There were 1003 participants (501 in the intervention group vs. 502 control group). The detailed sample size calculations, assumptions and eligibility for study sites were published in the main trial protocol [9].

### Study variables

The study captured key variables known to influence outcomes of TBI in accordance with the National Institute of Neurological Disorders and Stroke, and orthopaedic injuries, aligned with the global surgery agenda to enhance national surgical care plans [13], [14]. These variables included history of head trauma, head CT findings, intraoperative details, definitive surgical treatment, demographic factors, comorbidities, commute distance, injury mechanisms, road user categories, helmet use, prehospital care and transport, referral intervals, multiplicity of injuries, and injury severity as assessed by the Glasgow Coma Scale (GCS) and Kampala Trauma Score (KTS) [15].

The outcome variables were all-cause 90-day mortality and morbidity related TBI and musculoskeletal injuries. Mortality was defined as the death rate within the population. TBI morbidity was evaluated using the 5-point Glasgow Outcome Scale (GOS), while musculoskeletal morbidity was measured through the Trauma Outcome Measure Score (TOMS) at 90 days, compared to the Trauma Expectation Factor Score (TEFS) at admission. These instruments gauge patients’ recovery concerning pain, physical function, disability, satisfaction, and overall quality of life [16], [17].

### Statistical analysis

Descriptive statistics were employed to analyse participants’ baseline characteristics. Following established methodologies, the GOS was categorized into unfavourable (scores 1-3) and favourable (scores 4-5) outcomes [16]. Musculoskeletal injury outcomes were similarly classified as unfavourable (TOMS < TEFS) and favourable (TOMS ≥ TEFS) [17]. Mixed-effects logistic regression models, specifically the “melogit” command, were used to identify factors related to 90-day mortality and unfavourable morbidity outcomes, while adjusting for confounding variables. This approach accounted for baseline variations, intervention effects, and potential imbalances due to loss to follow-up, assuming that missing data were random.

The treatment allocation (intervention vs. control) served as the fixed effect unit of analysis. Odds ratios (ORs) and 95% confidence intervals (CIs) were reported as effect size estimates. The intra-cluster correlation coefficient (ICC) was calculated using the “estat icc” command in the mixed-effects restricted maximum likelihood (REML) regression model, as the ratio of between-cluster outcome variance to total outcome variance between and within clusters.

Covariates were assessed for confounding and effect modification using Cochran-Mantel-Haenszel statistics. Statistically significant variables in bivariate analysis, but not in Mantel-Haenszel strata, were excluded from the multivariable model. Relevant random variables with p<0.2 and those with biological plausibility such as KTS and GCS were retained. Analyses were conducted in Stata 15.0 at a 95% confidence interval, with significance at *p*<0.05.

### Ethical consideration

The Uganda National Council for Science and Technology approved the study (Ref: SS 5082), and all participants or their legally authorised representatives gave written informed consent before enrolment.

## RESULTS

The sociodemographic and clinical characteristics of study participants are detailed in the main trial report [preprint]. Briefly, of the 1003 participants, 82% (n=817) were male and 18.5% (n=186) were female, with a median age of 28 years (IQR: 22-37 years). Musculoskeletal injuries were present in 79.7% (799/1003), and head trauma in 94.6% (949/1003). Only 25.3% (254/1003) used helmets, and symptomatic TBI was present in 69.7% (699/1003). The overall median KTS and GCS were 8 (IQR: 7-9), and 14 (IQR: 11-15) respectively.

### Factors associated with 90-day all-cause mortality

Bivariate analyses of factors associated with 90-day all-cause mortality is presented in (Table 1). Of the 1003 participants, 88.4% (n=887) had complete 90-day follow-up data, of whom 9.2% (n=82/887) died. Variables associated with mortality included mode of arrival, multiplicity of injuries, injury severity based on KTS and GCS, head and brain CT diagnosis, and type of neurosurgical intervention (Table 1).

**Table 1:**
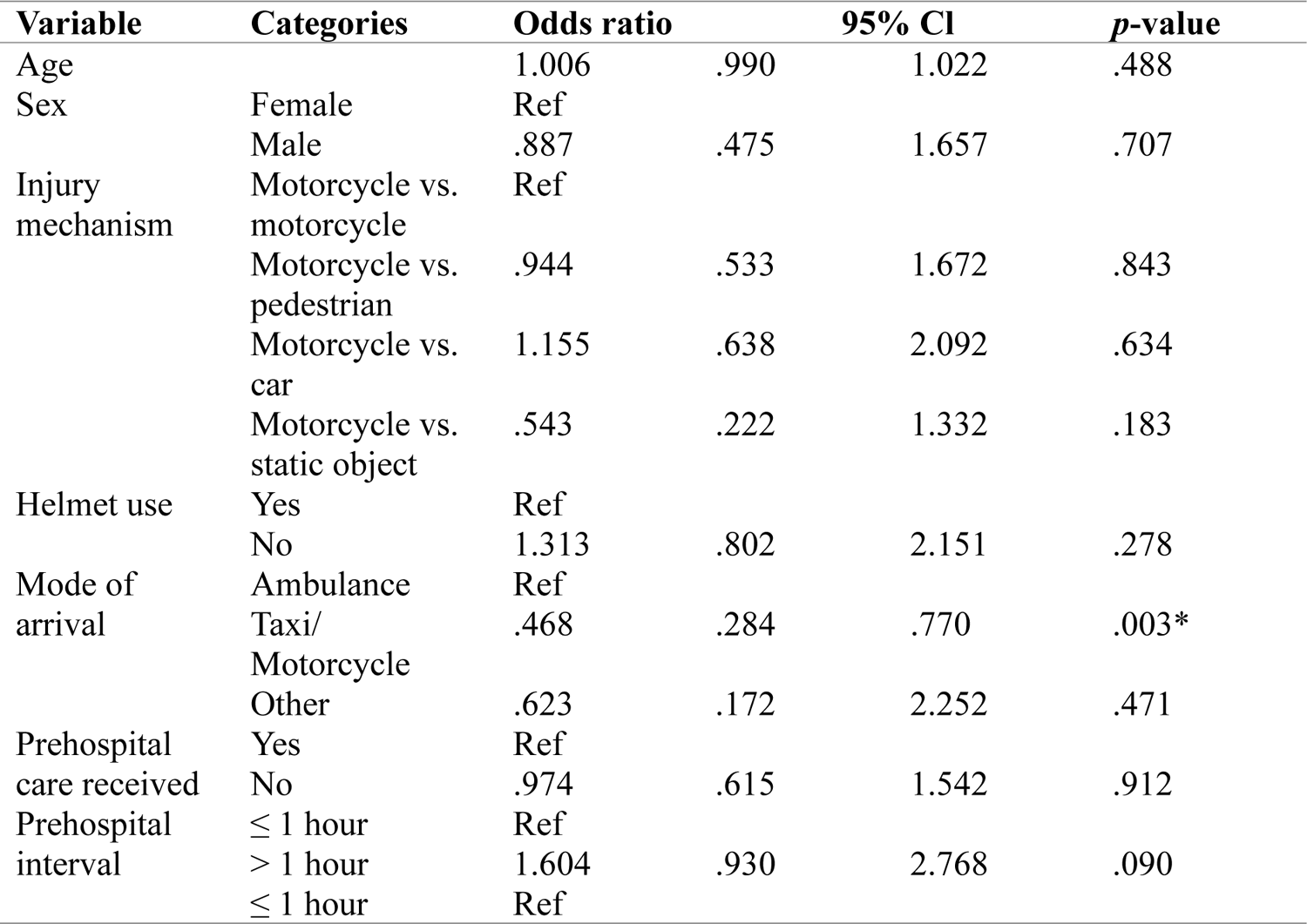
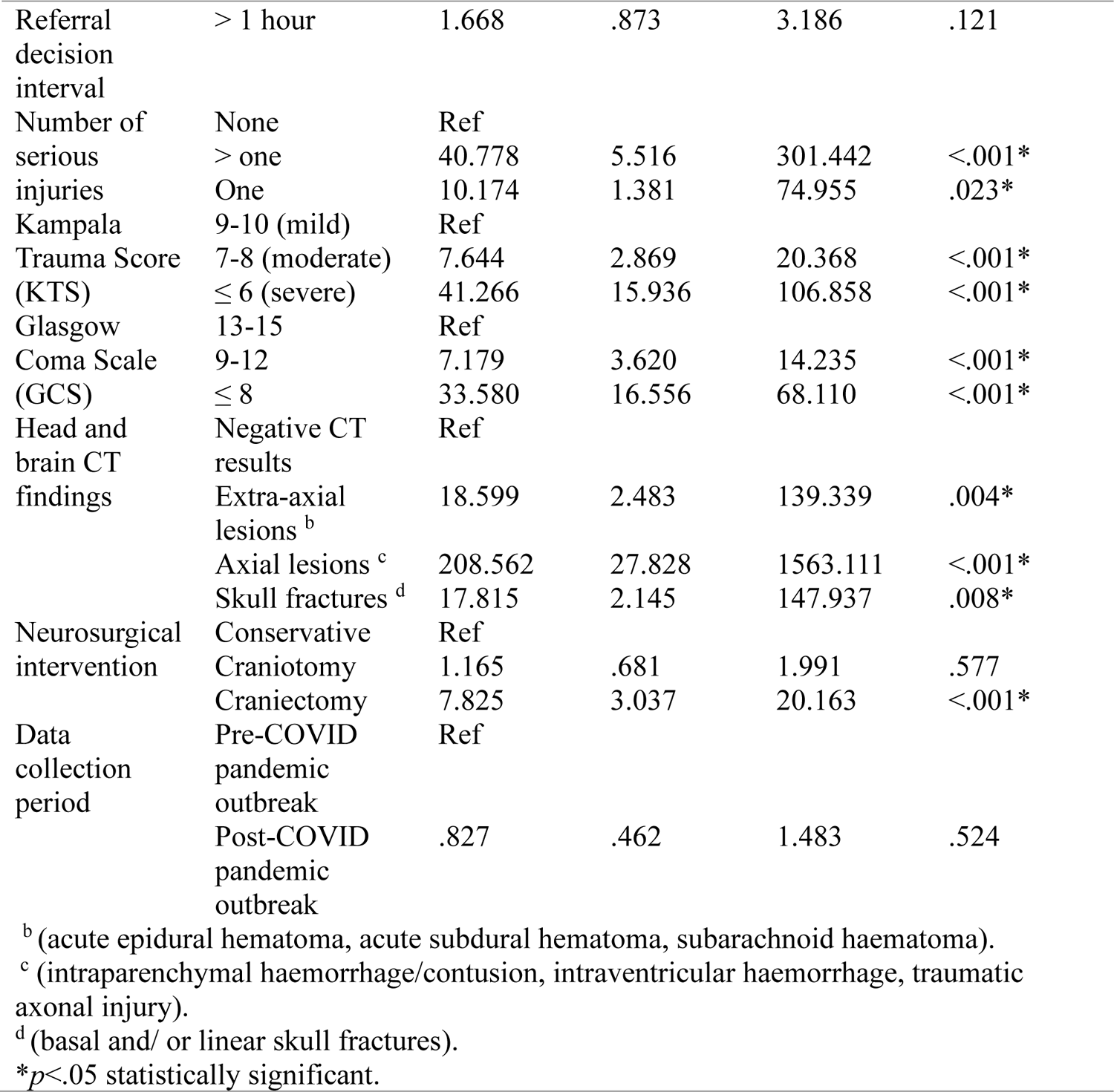
Bivariate analyses of factors associated with all-cause 90-day mortality.

Multivariable mixed-effects regression analysis, adjusting for confounding, identified several statistically significant predictors of 90-day all-cause mortality (Table 2). These included: referral to hospital dispatch interval >1 hour [OR 4.215, 95% CI (1.802-9.858)], KTS ≤6 [OR 7.696, 95% CI (1.932-30.653)], GCS 9-12 [OR 3.432, 95% CI (1.194-9.870)], and GCS ≤8 [OR 6.919, 95% CI (2.212-21.645)]. Compared to participants with negative CT scans, the highest odds of mortality were among those with intra-axial lesions [OR 78.647, 95% CI (9.871-626.587)], followed by extra-axial lesions [OR 11.933, 95% CI (1.386-102.750)] and skull fractures [OR 11.366, 95% CI (1.197-107.977)]. Conversely, decompressive craniotomy was associated with lower odds of 90-day mortality [OR .260, 95% CI (.0954-.706)] (Table 2).

**Table 2:** Multivariable analyses of factors associated with all-cause 90-day mortality.

| Variable | Categories | Adjusted Odds ratio | 95% CI |  | p-value |
| --- | --- | --- | --- | --- | --- |
|  |  |  | Lower | Upper |  |
| Prehospital care received | Yes | Ref |  |  |  |
|  | No | 1.277 | .659 | 2.473 | .468 |
| Referral decision to discharge interval | ≤ 1 hour (Ref) |  |  |  |  |
|  | > 1 hour | 4.215 | 1.802 | 9.858 | .001* |
| Kampala Trauma Score (KTS) | (9-10) | Ref |  |  |  |
|  | (7-8) | 2.670 | .737 | 9.678 | .135 |
|  | (≤ 6) | 7.696 | 1.932 | 30.653 | .004* |
| Glasgow coma score (GCS) | 13-15 | Ref |  |  |  |
|  | 9-12 | 3.432 | 1.194 | 9.870 | .022* |
|  | ≤ 8 | 6.919 | 2.212 | 21.645 | .001* |
| Head and brain CT diagnosis | Negative CT results | Ref |  |  |  |
|  | Extra-axial lesions <sup>b</sup> | 11.933 | 1.386 | 102.750 | .024* |
|  | Axial lesions <sup>c</sup> | 78.647 | 9.871 | 626.587 | <.001* |
|  | Skull fracture <sup>d</sup> | 11.366 | 1.197 | 107.977 | .034* |
| Neurosurgical treatment | Conservative | Ref |  |  |  |
|  | Decompressive Craniotomy | .260 | .095 | .706 | .008* |
|  | Decompressive craniectomy | .585 | .156 | 2.189 | .426 |

\*p<.05 statistically significant, Residual ICC =.0357, 95% CI (.002-.453) mixed-effects model.
<sup>b</sup> (acute epidural hematoma, acute subdural hematoma, subarachnoid haematoma).
<sup>c</sup> (intraparenchymal haemorrhage/contusion, intraventricular haemorrhage, traumatic axonal injury).
<sup>d</sup> (basal and/ or linear skull fractures).

### Factors associated with morbidity of neurological injuries

Table 3 summarises the bivariate analysis of factors associated with unfavourable Glasgow Outcome Scale (GOS) scores. The median GOS was 5 (IQR: 4-5), and the proportion of participants with unfavourable GOS (scores 1-3) was 14.5% (129/887). Statistically significant factors associated with unfavourable GOS included age, marital status, mode of prehospital transportation, prehospital interval, number of injuries, injury severity based on KTS and GCS, head and brain CT findings, neurosurgical treatment, and presence of comorbidities.

**Table 3:** Bivariate analysis of factors associated with unfavourable Glasgow outcome scale.

| Variable | Categories | Odds ratio | 95% Conf. Interval |  | P-value |
| --- | --- | --- | --- | --- | --- |
| Age |  | 1.023 | 1.010 | 1.037 | <.001* |
| Sex | Female | Ref |  |  |  |
|  | Male | 1.314 | .821 | 2.104 | .255 |
| Marital status | Single | Ref |  |  |  |
|  | Married | 1.610 | 1.053 | 2.463 | .028* |
|  | Divorced/separated | 4.832 | 2.119 | 9.611 | <.001* |
| Injury mechanism | Motorcycle-motorcycle | Ref |  |  |  |
|  | Motorcycle-pedestrian | .985 | .609 | 1.591 | .949 |
|  | Motorcycle-car | 1.418 | .874 | 2.301 | .157 |
|  | Motorcycle-static object | .672 | .336 | 1.343 | .260 |
| Helmet use | Yes | Ref |  |  |  |
|  | No | 1.164 | .768 | 1.763 | .475 |
| Mode of arrival | Ambulance | Ref |  |  |  |
|  | Taxi/Motorcycle | .584 | .394 | .865 | .007* |
| Prehospital care received | Yes | Ref |  |  |  |
|  | No | .983 | .673 | 1.436 | .929 |
| Prehospital interval | ≤ 1 hour | Ref |  |  |  |
|  | > 1 hour | 1.645 | 1.058 | 2.559 | .027* |
| Referral exit interval | ≤ 1 hour | Ref |  |  |  |
|  | > 1 hour | 1.175 | .724 | 1.908 | .515 |
| Number of serious injuries |  | .191 | .131 | .277 | <.001* |
| Kampala Trauma Score (KTS) |  | 4.870 | 3.617 | 6.555 | <.001* |
| Glasgow Coma Scale (GCS) |  | 4.512 | 3.425 | 5.946 | <.001* |
| Head and brain CT findings |  | 1.023 | .903 | 1.160 | .718 |
| Neurosurgical treatment |  | 2.237 | 1.608 | 3.111 | <.001* |
| Comorbidities | No | Ref |  |  |  |
|  | Yes | 2.811 | 1.501 | 5.267 | <.001* |
| Recruitment period | Pre-COVID pandemic outbreak | Ref |  |  |  |
|  | Post-COVID pandemic outbreak | .771 | .478 | 1.245 | .287 |

\**p*<.05 statistically significant

Multivariable mixed-effects regression analysis, adjusting for confounding, identified several key factors associated with unfavourable GOS scores (Table 4). These included: increasing age [OR 1.029, 95% CI (1.013-1.045)], sustaining more than one injury [OR 4.559, 95% CI (1.185-17.531)], KTS 7-8 [OR 2.755, 95% CI (1.285-5.906)], KTS ≤6 [OR 7.551, 95% CI (2.815-20.255)], GCS 9-12 [OR 4.071, 95% CI (1.901-8.719)], and GCS ≤8 [OR 13.779, 95% CI (5.643-33.645)]. Conversely, craniotomy was associated with reduced odds of unfavourable GOS [OR .490, 95% CI (.075-.295)] (Table 4).

**Table 4:** Multivariable analyses of factors associated with unfavourable Glasgow outcome scale.

| Variable | Categories | Adjusted Odds ratio | 95% CI |  | p-value |
| --- | --- | --- | --- | --- | --- |
|  |  |  | Lower | Upper |  |
| Age |  | 1.029 | 1.013 | 1.045 | <.001* |
| Serious injuries | None | Ref |  |  |  |
|  | > one | 4.559 | 1.185 | 17.531 | .027* |
|  | One | 2.327 | .662 | 8.182 | .188 |
| Kampala Trauma Score (KTS) | (9-10) | Ref |  |  |  |
|  | (7-8) | 2.755 | 1.285 | 5.906 | .009* |
| | ($\leq 6$) | 7.551 | 2.815 | 20.255 | <.001* |
| Glasgow coma score (GCS) | 13-15 | Ref |  |  |  |
|  | 9-12 | 4.071 | 1.901 | 8.719 | <.001* |
| | $\leq 8$ | 13.779 | 5.643 | 33.645 | <.001* |
| Neurosurgical treatment | Watchful waiting | Ref |  |  |  |
|  | Decompressive craniotomy | .149 | .075 | .295 | <.001* |
|  | Decompressive craniectomy | .794 | .244 | 2.580 | .701 |

\*p<.05 statistically significant, Residual ICC =.129, 95% CI (.018-.548) mixed-effects model.

### Factors associated with morbidity of musculoskeletal injuries

Among the 637 of 799 (79.7%) patients with musculoskeletal injuries who had complete Trauma Expectations Factor (TEFS) and Trauma Outcome Measure (TOMS) scores at 90 days, the majority (75.0%, 478/637) had a favourable outcome, with trauma outcome scores equal to or exceeding their trauma expectation scores. As shown in Table 5, factors associated with unfavourable TOMS scores in the bivariate analysis included: increasing age, being married and presence of comorbidities.

**Table 5:**
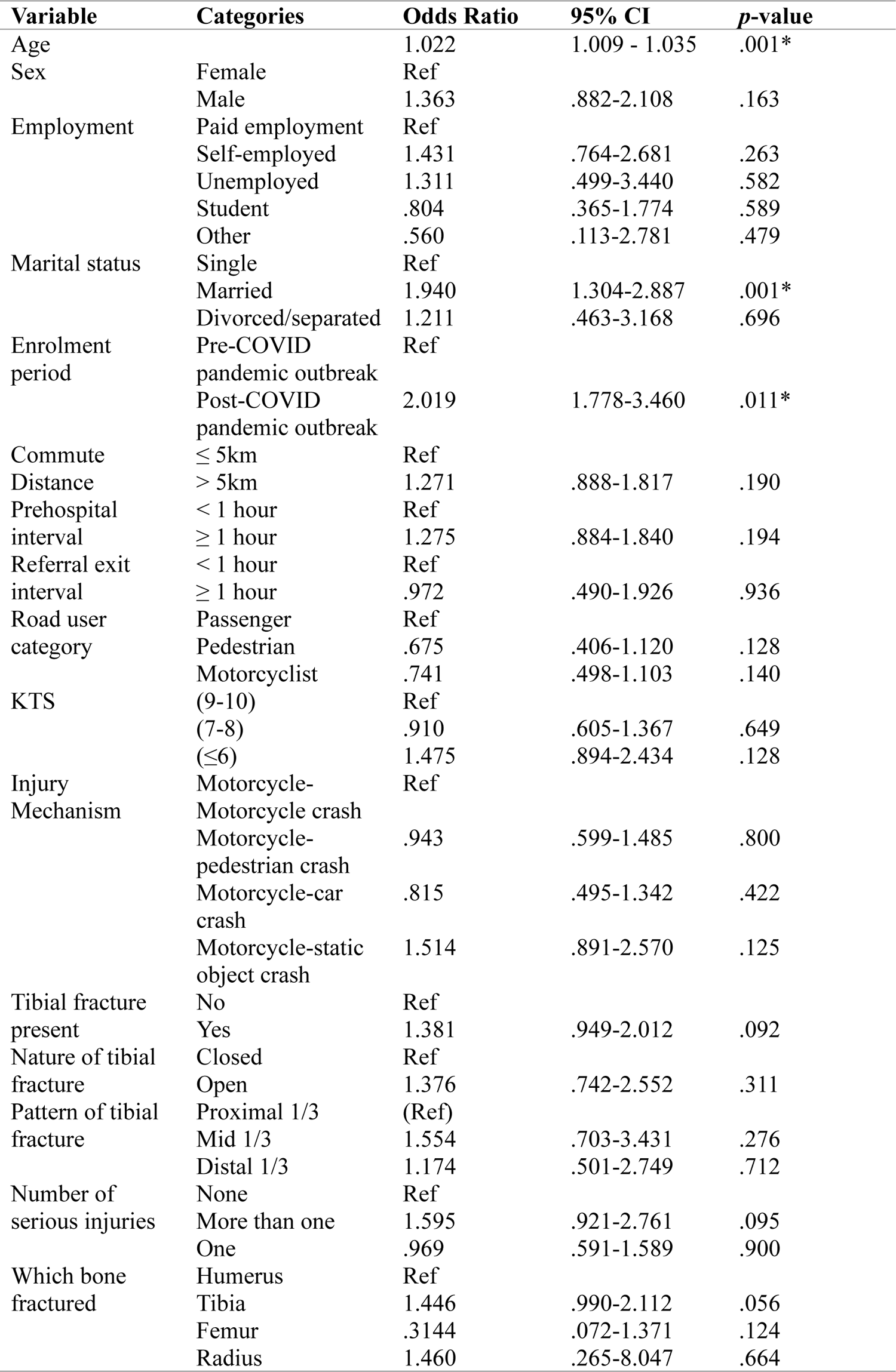
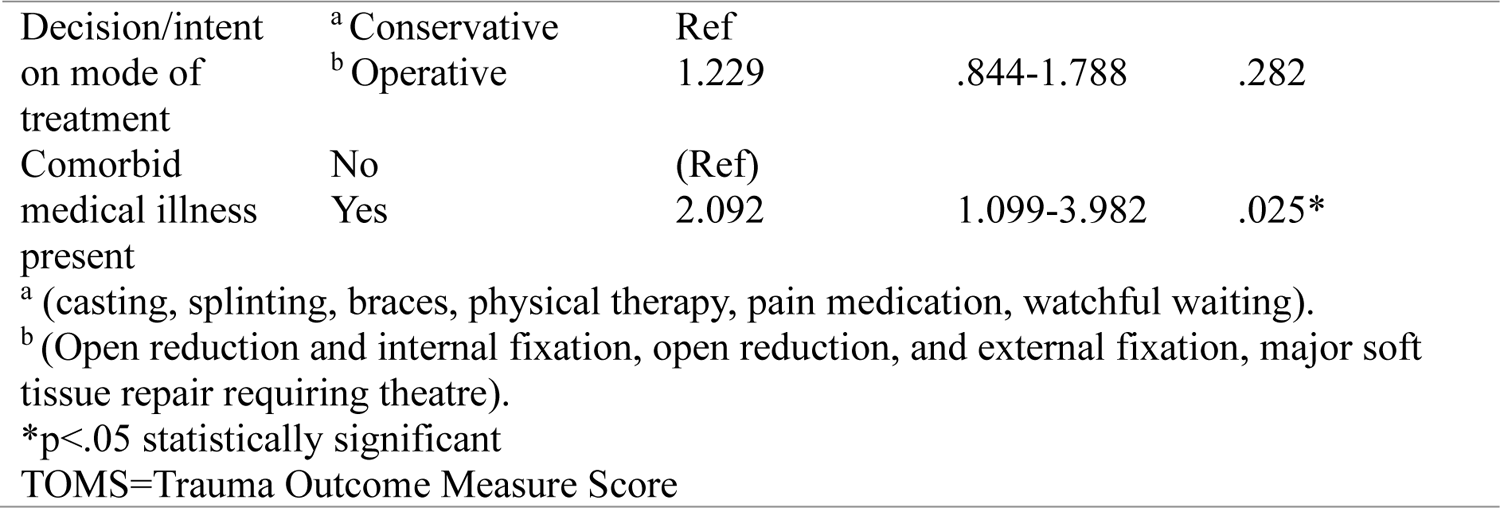
Bivariate analysis of factors associated with unfavourable TOMS for musculoskeletal injuries.

Multivariable mixed-effects logistic regression analysis of factors associated with unfavourable patient-reported outcome measures (PROMs) for musculoskeletal injuries is summarized in Table 6. The results indicate that being married [OR 1.984, 95% CI (1.322–2.976)], sustaining multiple injuries [OR 1.762, 95% CI (1.001–3.100)], and being enrolled after the onset of the COVID-19 pandemic [OR 2.095, 95% CI (1.199–3.659)] were associated with higher odds of unfavourable PROM scores.

**Table 6:**
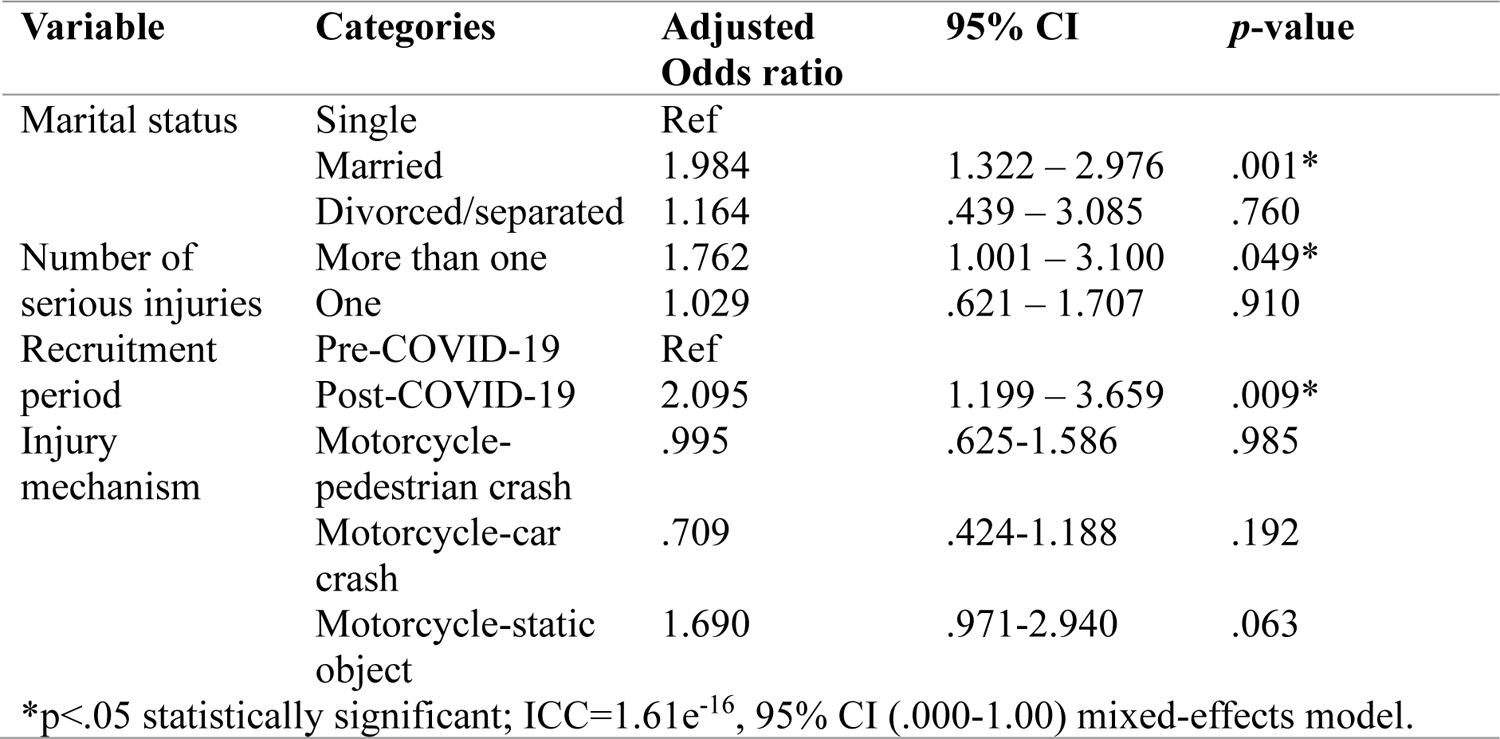
Multivariable mixed effects regression analysis of factors associated with unfavourable trauma outcome measure scores for musculoskeletal injuries.

| Variable | Categories | Adjusted Odds ratio | 95% CI | p-value |
| --- | --- | --- | --- | --- |
| Marital status | Single | Ref |  |  |
|  | Married | 1.984 | 1.322 – 2.976 | .001* |
|  | Divorced/separated | 1.164 | .439 – 3.085 | .760 |
| Number of serious injuries | More than one | 1.762 | 1.001 – 3.100 | .049* |
|  | One | 1.029 | .621 – 1.707 | .910 |
| Recruitment period | Pre-COVID-19 | Ref |  |  |
|  | Post-COVID-19 | 2.095 | 1.199 – 3.659 | .009* |
| Injury mechanism | Motorcycle-pedestrian crash | .995 | .625-1.586 | .985 |
|  | Motorcycle-car crash | .709 | .424-1.188 | .192 |
|  | Motorcycle-static object | 1.690 | .971-2.940 | .063 |

\*p<.05 statistically significant; ICC=1.61e<sup>-16</sup>, 95% CI (.000-1.00) mixed-effects model.

## Discussion

This study investigated factors associated with all-cause mortality and patient-related morbidity associated withto motorcycle crash injuries. It found that patients referred from a level III to level I trauma centre who experienced a delay of over one hour between referral decision and dispatch were four times more likely to die than those transferred within one hour. Corroborating findings from a Ugandan study found increased mortality for neurosurgical referrals arriving more than 4 hours post-injury [18], and our study has identified delayed referral decision execution as a critical contributor to these delays.

Consistent with a Dutch study of 22,525 moderately to severely injured patients [19], we observed no association between pre-hospital transportation time exceeding one hour and mortality. Evidence from systematic reviews indicate injury-intervention interval impact mortality for neurological and hemodynamically unstable orthopaedic injuries, while prehospital care is desirable over expedited transport for more stable trauma patients [20]. The present study expands the evidence base regarding the effect of prehospital time on mortality of time-dependent injuries in LMICs context, given most existing literature has emerged from HICs [20].

This study found helmet utilization at a mere 25%, insufficient to establish a statistically significant correlation with mortality and GOS outcomes. Notably, this rate exceeds the 20% usage documented in neighbouring Kenya but far below the global average [4].

We observed that injury severity based on KTS≤6 was nearly associated with eightfold mortality at 90-days. On the other hand, there was more than threefold and sevenfold increased mortality for participants with admission GCS score of 9-12 and ≤8, respectively. Both KTS [21] and GCS [22] have previously demonstrated the ability to predict mortality in multiply injured and TBI patients respectively. The TRACK-TBI cohort study of 1196 patients aged ≥17 years from 18 level I trauma centres found nearly four-fold increased mortality in those with GCS≤12 versus GCS 13-15 at 5-year follow-up [23]. Similarly, a retrospective cohort study from the American College of Surgeons’ National Trauma Data Bank identified admission GCS<9 as a strong predictor of mortality in motorcycle-related injuries [24]. In a Ugandan tertiary hospital cohort of 324 TBI patients, GCS score of <8 and 9-12 were associated with 6.6 and 2.4 increased risk of deaths respectively compared to GCS score of 13-15 [11].

In addition to mortality, we found both KTS and GCS predicted unfavourable GOS for functional independence. Participants with KTS 7-8 had 3 times the increase in the likelihood of worse GOS 1-3, rising to 7.5 times for KTS ≤6. Similarly, admission GCS score of 9-12 was associated with 4 times higher odds of unfavourable GOS at 90 days, increasing to 13-fold for GCS score of ≤8. Studies have found the GCS to be superior to the KTS in discriminating outcomes of TBI, although the primary focus has largely been on mortality rather than functional independence measures [25].

Our results demonstrate injury severity as the strongest predictor of TBI outcomes, corroborating with existing literature. In the USA TRACK-TBI study, only 72% of patients were functionally independent at 1-year post-TBI, with significantly lower odds of functional independence and complete recovery for patients with GCS score of 9-12 and <8 than in patients with GCS score of 13-15 [23]. However, evidence from the European CENTER-TBI study of 2376 subjects found even those with GCS score of 13-15 previously presumed to be mild injury severity had only 50% return to pre-injury functional independence by 6 months [26], obviating the need to contextualize trauma scores along with clinical evaluation, biomarkers, imaging and psychosocial-environmental modifiers.

We identified increasing age and the number of serious injuries per KTS as modifiers of adverse functional outcomes. Observational evidence from Norway corroborate young age ≤16 with favourable GOS while elderly ≥75 have worse prognosis [27]. Further, our study found participants with more than one acute injury had a 4.5-fold increase in the likelihood of unfavourable GOS. Emerging evidence demonstrate that TBI could enhance the healing of extra-cranial injuries such as fractures by modulating the immune system and bone metabolism [28]. However, multiplicity of injuries not only increases severity, but also imposes added stress from surgery and anaesthesia for associated injuries, elevating disability risk. The TRACK-TBI retrospective analysis of 1835 patients found those undergoing extra-cranial surgery and anaesthesia had worse functional outcomes at 6 months [29]. These findings underscore documenting extra-cranial injuries as essential for risk stratification to enhance trauma triage and registry protocols.

We found that, compared to individuals with negative head and brain CT scans, the presence of a skull fracture was associated with an 11-fold increased risk of 90-day mortality. This risk further escalated to 12 times in the presence of extra-axial lesions such as acute epidural, subdural or subarachnoid haemorrhages, and increased dramatically to 78 times if axial lesions such as contusions, intraventricular haemorrhage or traumatic axonal injury were detected. The study’s findings substantiate the hypothesis that surgically accessible extra-axial lesions are associated with lower mortality compared to more inaccessible axial lesions associated with cerebral oedema, elevated intracranial pressure, and brain herniation [30]. However, in agreement with Quach et al [31], the intra-axial/extra-axial classification failed to predict functional outcomes related to morbidity, as most patients with extra-axial lesions, including diffuse axial injuries, ultimately succumbed to death in our resource-limited environment.

While we observed decompressive craniotomy decreased 90-day mortality by 74.0% and unfavourable functional outcomes by 85.1%, the respective reductions were less pronounced at 41.5% and 20.6% for decompressive craniectomy relative to those managed conservatively. This discrepancy may stem from a larger proportion of our participants receiving craniotomy (23.5%) versus craniectomy (2.5%), as the majority of lesions were epidural (17.4%) rather than subdural hematomas (9.5%), but the results also conform to existing literature on the relative occurrence of intracranial haematomas, application, and benefits of the respective surgical approaches [11], [32]. Moreover, the results can also be interpreted so that decompressive craniectomy was performed in patients who had more severe and diffuse parenchymal intracranial injuries on head CT compared to patients treated with craniotomy for focal lesions, which may explain part of the difference. The optimal surgical management of intracranial haemorrhage has been contentious over the past decade with variable practice [33]. Proponents of decompressive craniectomy in which part of the skull bone flap is removed temporarily have emphasized its ability to control post-operative swelling and reduce intracranial pressure, which may otherwise be difficult to adequately manage in limited resource settings [33], [34]. The international RESCUE-ASDH randomized trial of 228 acute subdural hematoma patients found no difference in 6-month functional independence between craniotomy and decompressive craniectomy, although the craniectomy group trended towards higher one-year mortality, vegetative state, and wound infection rates [32]. The 24-month results of the RESCUEicp study showed that decompressive craniectomy significantly reduces the mortality rate in patients with traumatic intracranial hypertension compared to standard medical treatment. In addition, a higher percentage of patients in the surgical group showed functional improvement over time compared to patients treated medically alone, indicating a sustained survival benefit and some functional recovery [35].

A comprehensive systematic review and meta-analysis of 4,269 subdural hematoma patients found more favourable outcomes in the craniotomy group in terms of mortality, functional independence, and reoperation rates, compared to decompressive craniectomy [36]. However, craniectomy was associated with lower likelihood of residual hematomas. Moreover, the considerably worse outcomes observed with decompressive craniectomy were linked to worse baseline pre-operative injury characteristics in the patients requiring more aggressive surgical intervention [36]. Another meta-analysis of 6,848 patients found craniotomy was associated with a 20% reduction in mortality rates and 1.3 times higher likelihood of good functional outcomes compared to decompressive craniectomy [33]. However, the authors recommended a tailored, patient-centred approach to decision-making, since other factors such as extra-cranial injuries, concurrent epidural hematoma, and intracranial pressure also influenced the outcomes [33].

Based on the evidence from the CENTER-TBI study, patients with GCS < 12 and isolated hematomas ≥ 30 cc show better functional outcomes when treated surgically whereas those with GCS>12 and hematomas < 30 cc do well when managed conservatively [37]. The 2020 brain trauma foundation guidelines recommend a large decompressive craniectomy for late refractory intracranial pressure (ICP) elevation to reduce ICP, mortality and duration of intensive care, though favourable functional recovery may not be guaranteed [38]. Thus, the existing evidence suggests that optimal surgical approach should be considered in the context of the individual patient’s unique clinical presentation and injury profile which provides helpful context for interpreting the discrepancies seen in our own findings between the two surgical approaches,but identifying the ideal patient profile who would benefit the most from decompressive craniectomy should be the subject of future research.

After adjusting for confounding variables, factors associated with unfavourable patient-reported trauma outcomes for musculoskeletal injuries were multiple injuries [OR 1.762], being married [OR 1.983], and sustaining the injury during the post-COVID-19 pandemic period [OR 2.095]. Comorbid lower extremity injuries are associated with higher injury severity and poorer health-related quality of life following major trauma [39]. While physical function and pain improve over time, social health lags behind, especially for lower extremity trauma [40]. Failure to meet psychosocial demands and family commitments, such as marital and financial responsibilities, can contribute to unfavourable patient-reported outcomes among married individuals. The socioeconomic impact of the COVID-19 pandemic appears to have exacerbated this phenomenon in family-oriented participants. These findings underscore the need for patient-centred, injury-specific approaches during rehabilitation to promote full recovery, as definitions of good outcomes vary across cultural and family contexts. Thus, psychosocial patient-reported outcomes should be included as indicators when evaluating functional musculoskeletal care systems to better assess return to pre-injury status.

### Study limitations

This study was not without limitations. First, the smaller numbers of some injuries made analyses of individual lesions statistically unsound, and aggregating TBI and dichotomizing trauma outcomes could have resulted in loss of information. The use of predictive models and neural networks with machine learning algorithms is an emerging approach to addressing this challenge.

Secondly, 90-day follow-up was considered short-term, as functional improvements may not be detected by the Glasgow Outcome Scale until 6-12 months post-injury, moreover mortality may occur beyond this period [27].

Thirdly, the study population was limited to motorcycle crash-related injuries, so the results may not generalize to other injury mechanisms such as penetrating TBI which present distinct outcome characteristics. Moreover, the study was restricted to clinical assessment and head CT, excluding prognostically informative MRI and blood biomarkers.

Lastly, 11.6% of participants had missing data on outcome variables which may introduce bias following exclusion from the analysis. Additionally, the potential for interobserver variations in data acquisition could not be ruled out.

Nevertheless, this is one of the largest studies to evaluate mortality and patient-reported morbidity outcomes of neurological and musculoskeletal injuries unique to motorcycle crashes in Uganda. Moreover, our analyses were robust given risk adjustment, and all experiments were reviewed and approved by authors of the parent trial.

## Conclusions

The research findings indicate that several key factors were determinants of 90-day mortality and unfavourable outcome, including referral decision to dispatch interval, presence of axial and extra-axial lesions, skull fractures, multiplicity of injuries, and injury severity based on the KTS and GCS. Conversely, decompressive craniotomy was identified as a protective factor. For patient-reported outcomes related to musculoskeletal injuries, the key determinants were multiplicity of injuries, marital status, and sustaining injuries during the post-pandemic period. These critical factors should be incorporated into future prospective trials, trauma registries, and predictive machine learning models that incorporate demographic, injury mechanism, clinical, and CT imaging data. Additionally, long-term trials evaluating similar outcomes for diverse injury mechanisms such as motor vehicle crashes, falls, and assaults should be conducted.

## Supporting information

Ethical approval and data analysis codes

## Data Availability

All data produced are available online at https://data.mendeley.com/datasets/bgpmkpcwdt/1

The anonymized data underlying the findings of this ancillary study are available within this article. Additionally, the dataset and analysis code pertaining to both the ancillary study and the parent trial, which is registered with the Pan African Clinical Trial Registry (PACTR202308851460352), are publicly accessible [41].

## Acknowledgements

The authors express gratitude to the staff at medical referral institutions in Uganda, including Jinja, Mbarara, Hoima, Mubende, Kiryandongo, and Kampala International University Teaching Hospitals, for providing facilities that enabled data collection for this study. Additionally, the authors thank the study participants for dedicating their time to take part in the investigation.

## Author contributions

HL was the lead researcher who conceived, designed, and implemented the study, and wrote the initial draft of the manuscript. RS, PK, AK, BO and HML assisted with administrative tasks, data acquisition, and participant follow-up, and provided critical review of the manuscript. HL, MM, and AA were responsible for data analysis and cross-validation. BT critiqued the study methods. Funding for the project was obtained by JPP, MLW, BT and HL. MLW and JPP supervised the study and provided critical review of the final manuscript. All authors read and approved the final version of the manuscript.

## Patient consent for publication

Not applicable

## Ethical approval

Ethical approval for this research involving human participants was granted by the Research and Ethics Committees of Mbarara University of Science and Technology (Ref: 1/7; 05/5-9) and the Uganda National Council for Science and Technology (Ref: SS 5082).

## Provenance and peer review

Not commissioned, pending peer review

## List of abbreviations

GCS: Glasgow Coma Score
GOS: Glasgow Outcome Scale
GOSE: Glasgow Outcome Scale Extended
KTS: Kampala Trauma Score
LMICs: Low-Middle-Income Countries
HICS: High-Income Countries
CENTER-TBI: Collaborative European Neuro Trauma Effective Research in Traumatic Brain Injury
TRACK-TBI: Transforming Research and Clinical Knowledge in Traumatic Brain Injury
STITCH: Surgical Trial in Traumatic Intracerebral Haemorrhage

